# Advancing data science in drug development through an innovative computational framework for data sharing and statistical analysis

**DOI:** 10.1101/2021.02.16.21251799

**Authors:** Ann-Marie Mallon, Dieter A. Häring, Frank Dahlke, Piet Aarden, Soroosh Afyouni, Daniel Delbarre, Khaled El Emam, Habib Ganjgahi, Stephen Gardiner, Chun Hei Kwok, Dominique M. West, Ewan Straiton, Sibylle Haemmerle, Adam Huffman, Tom Hofmann, Luke J. Kelly, Peter Krusche, Marie-Claude Laramee, Karine Lheritier, Greg Ligozio, Aimee Readie, Luis Santos, Thomas E. Nichols, Janice Branson, Chris Holmes

## Abstract

**Background:** Novartis and the University of Oxford’s Big Data Institute (BDI) have established a research alliance with the aim to improve health care and drug development by making it more efficient and targeted. Using a combination of the latest statistical machine learning technology with an innovative IT platform developed to manage large volumes of anonymised data from numerous data sources and types we plan to identify novel patterns with clinical relevance which cannot be detected by humans alone to identify phenotypes and early predictors of patient disease activity and progression.

**Method:** The collaboration focuses on highly complex autoimmune diseases and develops a computational framework to assemble a research-ready dataset across numerous modalities. For the Multiple Sclerosis (MS) project, the collaboration has anonymised and integrated phase II to phase IV clinical and imaging trial data from ≈35,000 patients across all clinical phenotypes and collected in more than 2,200 centres worldwide. For the “IL-17” project, the collaboration has anonymised and integrated clinical and imaging data from over 30 phase II and III *Cosentyx* clinical trials including more than 15,000 patients, suffering from four autoimmune disorders (Psoriasis, Axial Spondyloarthritis, Psoriatic arthritis (PsA) and Rheumatoid arthritis (RA)).

**Results:** A fundamental component of successful data analysis and the collaborative development of novel machine learning methods on these rich data sets has been the construction of a research informatics framework that can capture the data at regular intervals where images could be anonymised and integrated with the de-identified clinical data, quality controlled and compiled into a research-ready relational database which would then be available to multi-disciplinary analysts. The collaborative development from a group of software developers, data wranglers, statisticians, clinicians, and domain scientists across both organisations has been key. This framework is innovative, as it facilitates collaborative data management and makes a complicated clinical trial data set from a pharmaceutical company available to academic researchers who become associated with the project.

**Conclusions:** An informatics framework has been developed to capture clinical trial data into a pipeline of anonymisation, quality control, data exploration, and subsequent integration into a database. Establishing this framework has been integral to the development of analytical tools.

## Background

Novartis and the University of Oxford’s Big Data Institute (BDI) have established a research alliance with the aim to improve health care and drug development by making it more efficient and targeted. Working with multi-disciplinary teams and using artificial intelligence (AI) and advanced analytics, the collaboration expects to transform how multiple large multidimensional (clinical, imaging, omics, biomarkers) datasets are combined, analysed and interpreted to identify phenotypes and early predictors of patient disease activity and progression, and to improve prognosis for patients. The collaboration currently focuses on two projects; one on Multiple Sclerosis (MS) and one on several autoimmune diseases treated with the -IL-17 antibody Cosentyx (secukinumab). The collaboration will also be making use of anonymised data from approximately 5 million patients from both the UK and international partner organisations, together with anonymised data captured from relevant Novartis clinical trials -in a total of approximately 50,000 patients. Using the BDI’s latest statistical machine learning technology and experience in data analysis, combined with Novartis’ clinical expertise and high-quality clinical trial data, the collaboration expects to better understand the underlying disease and early predictors of disease activity to improve prognosis for patients.

Here we describe the data anonymisation and the development of an innovative IT environment and AI technology, through which the alliance is working collaboratively to identify patterns, often across multiple data sources and types, which cannot be detected by humans alone. Novartis and the BDI expect to gain insights into the characteristics of specific, complex diseases and their pathways to understand what drives disease progression, and to understand commonalities between diseases.

## Methods

### A large-scale high-dimensional dataset

The collaboration currently focuses on Multiple Sclerosis (MS) and other inflammatory diseases in dermatology and rheumatology which are major areas of drug development. A computational framework and data management process have been established to facilitate the collaboration and enabling all data scientists to work together on the data. Throughout the next section we will describe the breadth and scope of the data contributed in both the Multiple Sclerosis and IL-17 projects, highlighting the variety of data types and modalities, the global nature of the data generation and the longitudinal aspects which increase the complexity of data management, curation and integration to enable us to produce research ready datasets of the highest utility to the statisticians and analysts.

### Clinical trial data from patients

The core data used in the collaboration stem from Novartis clinical trials. All trials were conducted in accordance with the provisions of the International Conference on Harmonisation guidelines for Good Clinical Practice and the principles of the Declaration of Helsinki. The trial populations were defined by eligibility criteria (i.e. inclusion and exclusion criteria), all trial procedures followed trial protocols, specifying the purpose of the experiment, the medical objectives, the endpoints, the assessments and assessment frequencies, as well as the statistical analysis methods to address the key-objectives of the trials. All trial protocols were approved by an institutional review board or ethics committee and all patients or their legal representatives gave written informed consent before any trial-related procedures were performed. In general, data from clinical trials can only be used if the usage is covered by the informed consent. Usage beyond the informed consent defined scope requires the data to be anonymised in order that they are no longer personal data. To maintain data privacy, all data have been anonymised before use for analyses by the collaboration. The collaboration uses dose finding studies (phase II), confirmatory clinical trials (phase III) which are designed to confirm the efficacy and safety of a new treatment option versus a control treatment to seek regulatory approval for the new treatment, and phase IIIb and IV studies which are typically open-label studies after the approval of a drug. For illustration purposes, the design of a typical confirmatory (phase III) clinical trial is presented in Figure 1.

**Figure 1:**
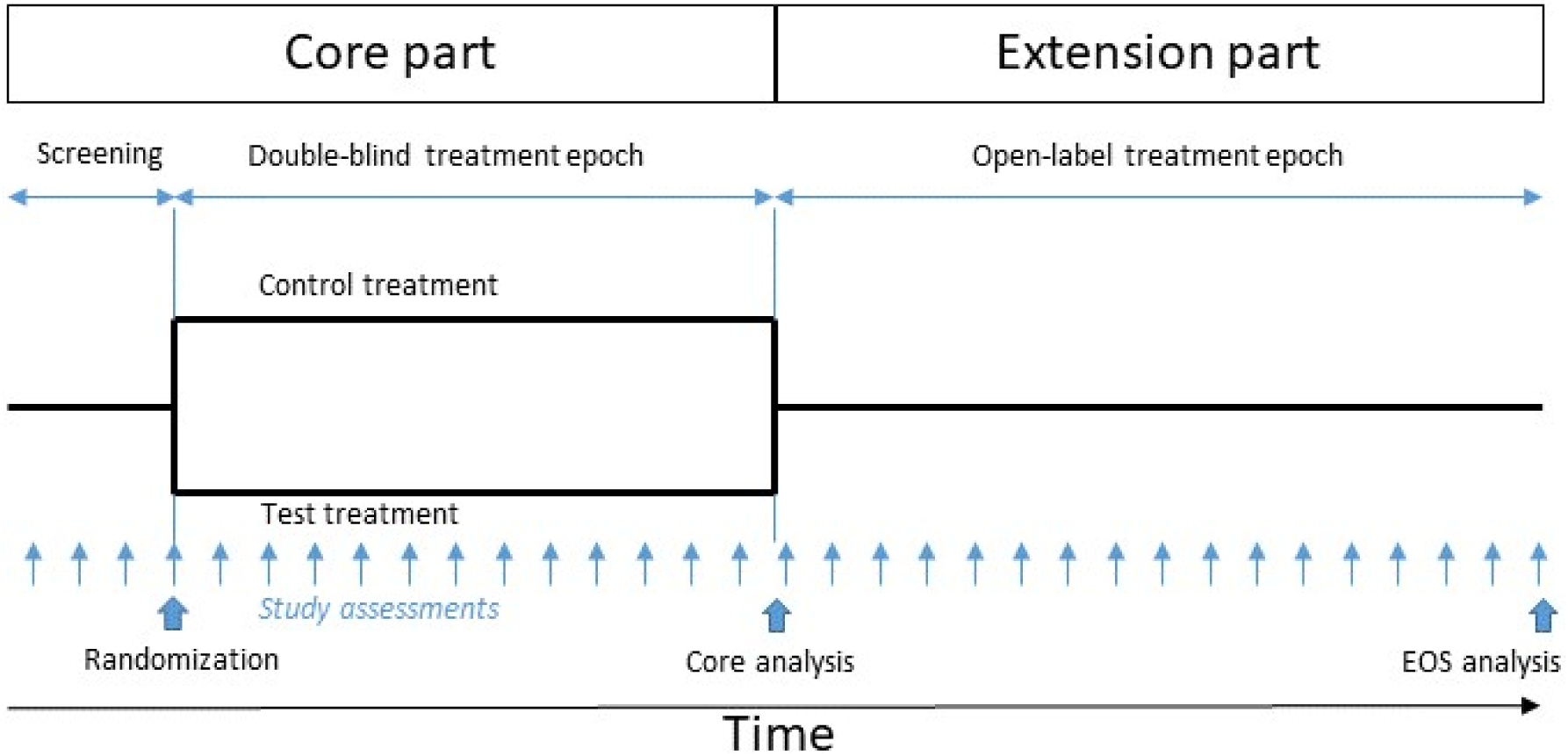
A schematic of a typical randomised clinical trial. A typical confirmatory phase III clinical trial, followed by an open-label extension study. During a screening period, and after signing informed consent, eligibility of the patient for the trial is assessed. Eligible patients may then be randomised to one or several test and control (placebo or active control) treatments. In a double-blind study, patients, physicians and study personnel are blinded to the patient’s treatment assignment until the core experiment has been completed and all assessments have been collected. Then the database is locked, the treatment allocation unblinded and the data analysed (core analysis). Often patients who complete a core study are offered to continue in an open-label study, for instance on the newly tested treatment until the new treatment option becomes available on the market. All assessments and study procedures are defined by a study protocol. EOS represents the end of the study.

### Multiple Sclerosis (MS) project

Multiple Sclerosis (MS) is a chronic, immune-mediated disease of the central nervous system (CNS) characterized by inflammation, demyelination, and axonal/neuronal destruction, ultimately leading to severe disability. MS is the most common autoimmune demyelinating disorder of the CNS, affecting approximately 2.3 million individuals worldwide [1].

MS typically affects young adults (mean age at onset 30 years) and women are affected more often than men. Reflecting the current understanding of MS, the disease course of MS can be grouped into 2 corresponding main MS categories [2]:

- relapsing MS (RMS): clinically isolated syndrome (CIS), relapsing-remitting MS (RRMS), active secondary progressive MS (SPMS)
- progressive MS: secondary progressive Multiple Sclerosis (SPMS) and primary progressive MS (PPMS)

For Multiple Sclerosis (MS), the collaboration has integrated data from a total of approximately 35,000 MS patients collected in more than 2,200 centres across 57 countries in 34 clinical trials (4 phase II trials of which 3 had extensions, 13 phase III trials of which 5 had extensions and 9 phase IV trials) which were conducted between years 2003 and now (2019 and ongoing). Over 88,000 patient-years of data is on record with individual patients being followed up for up to 15 years. Studies include randomised controlled clinical trials from three major drug development programs including patients from placebo- and active control arms, but also open-label and real-world studies which used other disease modifying therapies, in all phenotypes of MS patients:

- Relapsing Remitting MS (N∼32,000)
- Primary- and secondary progressive MS (N >2,800)

The database covers the entire spectrum of MS phenotypes including paediatric (N=235), treatment naive MS patients (N=5,445), but also patients who had the disease for >25 and up to 50 years (N=1,624).

The dataset also includes a wealth of different data modalities, including detailed information on demography and baseline characteristics, medical history, MS relapses (>16,000 relapses recorded) with symptoms and recovery grades, physical disability assessments (> 238,000 neurological assessments of the Expanded Disability Status Scale [EDSS]) covering all levels of disability (from EDSS=0 normal neurological exam with no disability to EDSS=10 death due to MS), 25-foot walking test (walking ability), 9-hole peg test (hand coordination), cognitive assessments (PASAT, SDMT), laboratory values, ECGs, vital signs, concomitant medication, detailed treatment information, and adverse event data. More than 13,000 patients have clinical records that also include MRI summary features. For >11,000 of these patients, we also have the raw MRI images available for analyses and feature extraction, totalling to >230,000 MRI scans, with T1-weighted (pre and post gadolinium contrast enhancement), T2-weighted, proton density, FLAIR, magnetization transfer, and diffusion-weighted sequences, longitudinal data being available for up to 12 years in individual patients. The collation and anonymization of these raw MRI images (see results) has been established in this collaboration to ensure they can be integrated into the overall research ready database.

### Interleukin-17 inhibitor project

*Cosentyx* (secukinumab) is a high-affinity recombinant, fully human monoclonal Interleukin-17A (IL-17A) antibody. By binding to human IL-17A, Cosentyx neutralizes the bioactivity of this cytokine. IL-17A is the central lymphokine of a defined subset of inflammatory T cells, which appear to be pivotal in several autoimmune and inflammatory processes. The collaboration has integrated data from a total of 16,576 randomised patients from over 21 phase II and 44 phase III *Cosentyx* clinical trials targeting four autoimmune disorders in dermatology and rheumatology. They are:

- Psoriasis (PsO)
- Axial spondyloarthritis (axSpA), including ankylosing spondylitis and non-radiographic axial spondyloarthritis
- Psoriatic arthritis (PsA)
- Rheumatoid arthritis (RA)

Each study records a range of measurements on each subject at multiple time points throughout the trial, with the duration of collection of patient data ranging from 12 weeks to 5 years. The dataset includes different data modalities ranging from demography and patient history to laboratory data and imaging. A number of assessments are collected as a standard in all clinical studies and are therefore available across indications, such as demography and baseline characteristics, medical history, ECGs, vital signs, concomitant medication, detailed treatment information, and adverse event data, Quality of Life questionnaires, and laboratory measurements from serum and whole blood samples. Genomic, proteomic, and transcriptomic data, as well as imaging data from MRI and X-ray scans are also available for some patients depending on the trial design. Overall, the diseases under study all have a commonality of inflammation in different regions of the body, therefore measurements of inflammation are a common assessment as well. Other datasets are collected in some of the indications due to common disease pattern. One such example is skin assessment, commonly done in PsO and PsA or joint assessments done in PsA, axSpA and RA. Additionally, MRI and X-ray images of affected body locations were taken. In axSpA, which manifests predominantly in the axial region, MRI and X-rays of the spine and sacroiliac joints are taken in order to monitor treatment response. In PsA, X-rays of the hands, wrists and feet are collected. This dataset will be used to model patient disease trajectories, as well as interpret and predict of multivariate longitudinal response to *Cosentyx*, in order to improve the clinical outcome of patients across the four autoimmune disorders.

## Results

A fundamental component for the successful data analysis and the collaborative development of novel machine learning methods on this rich data set has been the construction of an innovative research informatics framework that can capture the data at regular intervals from Novartis into a secure IT infrastructure at the BDI where data could be integrated, quality controlled and compiled in to a research ready relational database which would then be available to analysts. The development of this framework (Figure 2) in the BDI and the successful capture of the data (described previously in the Methods) into a versioned research ready dataset is described below.

**Figure 2:**
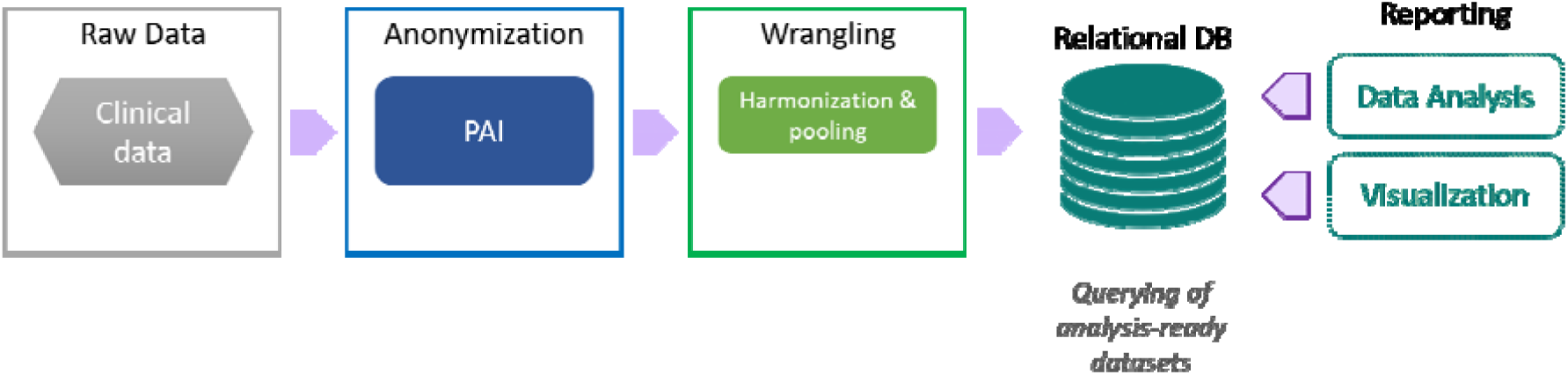
Innovative and robust informatics framework for high-dimensional clinical trial data. Raw clinical data is anonymised by Privacy Analytics, Inc. (PAI), followed by data wrangling that involves harmonisation and pooling of data. Data is then integrated into a relational database (DB) whereby users of the DB are able to obtain analysis-ready datasets through querying. Data within the DB can be used for data analysis and visualisation purposes.

### A secure and collaborative research infrastructure

A critical part of the research collaboration was to develop an IT infrastructure and corresponding information security architecture that was technologically feasible, business viable and foremost user desirable for the project. The design of the information security architecture and controls was governed by the need for the processing of a large amount of clinical data shared between two organisations. In order to design and implement proper information security, it was vital to understand the various stages in the process, the states of the data, the information security goals and the overall risks. After collecting and assessing all of this data, a custom information security architecture was defined and security controls and guidelines applied.

The overall results of the above methodology were that confidentiality was identified as the overall and most relevant information security goal within the project. This decision defined the fundamental principle that governed the design and implementation of the security controls.

1. Isolation from the existing Oxford BDI infrastructure whenever possible, at least for every activity involving non-anonymised data.
2. Encrypted data transfer via dedicated channels between both organisations to ensure confidentiality and integrity.
3. User access to the environment is proxied via a demilitarized zone (DMZ), which contains a certain set of jump hosts that serve as portals to the full environment.
4. Identity and access management is realized within the environment, providing authentication and authorisation services.
5. The log management is done at a central place, together with security information and event management.

Two separate environments were created for anonymisation and analytics work, and both of these were instantiated within a dedicated OpenStack private cloud, as two separate tenants. This ensures network, compute and storage isolation enforced at the hypervisor level. For data processing virtual clusters were created within each tenant, including an instance (virtual machine) with direct access to GPUs for accelerated work. The virtual resources within each tenant were defined and provisioned by means of Ansible roles and playbooks, for consistency and repeatability. Encrypted backups are made to an S3 object store. In short, we have produced a unique research computing infrastructure that provides high levels of security while providing a shared environment where both academic and industrial researchers can jointly work.

### Clinical Data Anonymisation

This section describes the anonymisation of the clinical trial data and the specific methods developed to anonymise MRI data to ensure data privacy.

#### Clinical Trial Data – Basic Principles

The process for anonymising the clinical trial data was intended to ensure that the risk of re-identifying participants in the dataset was below a pre-defined critical threshold. There are three key concepts in this risk-based anonymisation approach:

- The risk of re-identification can be measured quantitatively. Various models of adversaries and re-identification attacks have been developed and have demonstrated robustness in practice [3]. Metrics quantifying the probability of a successful re-identification have been developed based on these models. The specific metrics that we used are based on strict average risk models. These capture the average risk while ensuring that there are no population unique individuals in the anonymised dataset (i.e., in the context of the General Data Protection Regulation (GDPR), the likelihood of individuals being “singled out” is very small [4].
- The overall risk measurement takes into account the context of data processing as is illustrated in Figure 3. For example, if the anonymised dataset will be analysed in a secure compared to a less secure environment then less modification of the data is required to bring the risk of re-identifying a patient to below the targeted threshold. Checklists have been developed and validated to capture this context risk [3].

**Figure 3:**
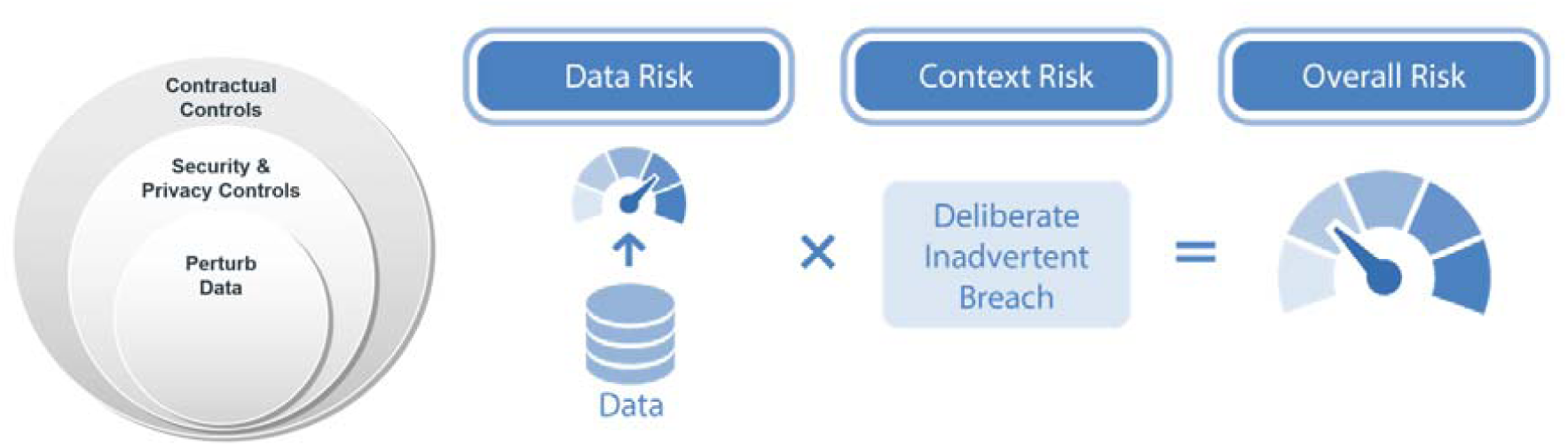
Clinical Trial Data Anonymisation. The overall risk of re-identification is a function of both the data risk and the context risk. The context risk is assessed by examining three re-identification attacks on a dataset: (a) a deliberate attack by an adversary, (b) an inadvertent re-identification by a data analyst where they recognize someone they know, and (c) a data breach occurring. The success of the three attacks is affected by the controls that are in place. The context consists of first the contractual controls which reduce the context risk. The residual risk is managed by security and privacy controls, which are also part of the context. The extent of these controls reduces the overall risk further. Then any residual risk is managed by perturbing or transforming the data.
- A specific threshold needs to be defined to determine what an acceptably low risk is. There are many precedents for what is deemed to be an acceptable threshold, including from regulators (see the review in [3]). The choice of a specific threshold from the precedent range takes into account the sensitivity of the data and the potential harm if there is a re-identification.

Once a threshold is defined and the re-identification risk is computed, taking into account the context, transformation may be required until the risk is below the defined threshold. The transformations can be performed to the data itself (e.g. by modifying variables that may lead to re-identification such as a patient’s age) or to the context (e.g. by modifying the security of the IT system). After each transformation the overall risk can be re-computed until it is below the threshold.

#### Justification for Threshold

The European Medicines Agency (EMA) has established a policy on the publication of clinical data for medicinal products [5] which requires applicants/sponsors to openly share clinical trial data. The guidelines accompanying the policy recommend a maximum risk threshold of 0.09. Health Canada implemented the same threshold for the sharing of clinical trial data [6]. This is the threshold that is used for the anonymization of the clinical data.

#### Calculation of Risk

The risk of re-identification is calculated only on the quasi-identifiers. The quasi-identifiers are variables that are knowable by an adversary. There are two general types of quasi-identifiers. The first are those which are in the public domain and can be collected from registries such as voter registration lists [7] and lien registries [8]. Examples of these include date of birth and ZIP/postal codes. The second are acquaintance quasi-identifiers, which are known by adversaries who are also acquaintances, such as neighbours, relatives, and co-workers. Acquaintance quasi-identifiers include the public ones as well as things like medical history and key events and dates. Once the quasi-identifiers are determined in a dataset, the probability of re-identification can be calculated.

The calculation of re-identification risk considers three potential attacks on the data, which we shall call T1, T2, and T3.

The first attack, T1, assumes that an adversary deliberately attempts to re-identify individuals in the dataset [9]. This means that the probability of re-identification is conditional on an attempted attack:

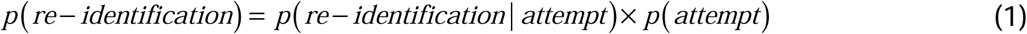

The first term captures the risk in the data and the second term captures the risk from the context. There are multiple estimators that can be used to evaluate data risk which vary in accuracy and scalability [3], [10]–[15].

Context risk has three components: security controls, privacy controls, and contractual controls. The strength of these controls as they were implemented at the BDI were assessed using a checklist. The checklist is reproduced elsewhere [3]. The responses to the checklist are converted into a conservative subjective probability. This means that the exact probability value is not known, but the modeled value is convincingly conservative (over estimates the context risk) but still allows us to model the controls that are in place and account for the benefits of stronger controls.

The premise of the controls for attack T1 is that the existence of stronger security controls (e.g., audit logs that are checked, analyst screening, and limited access), privacy controls (e.g., regular privacy training and a privacy officer), and contractual controls (e.g., all analysts have to sign a confidentiality agreement when working with the data) act as deterrents for an attempted attack and make it more difficult.

A T2 attack pertains to an inadvertent re-identification. This is when an analyst inadvertently or spontaneously recognizes someone that they know in the dataset as they are working on it. This type of risk is given by:

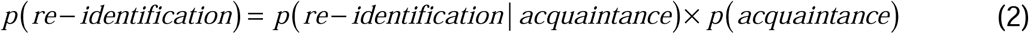

An inadvertent re-identification is contingent on an analyst knowing someone in the data. In our case this means that an analyst would know someone who has participated in a trial in this therapeutic area. This is estimated as: *p*(*acquaintance*) =1 − (1 − *v*)^150^ where *v* is the proportion of patients in the current studies compared to all studies in this therapeutic area over the same period and geography, which can be computed by gathering target recruitment data from https://clinicaltrials.gov/. The 150 value is the Dunbar number, which provides us with an estimate of the average number of individuals that an analyst would know. Dunbar’s has proven to be robust across multiple studies (for a literature review see [3]).

The third attack is when there is a data breach and the dataset is accessed by an adversary. This is modeled as follows:

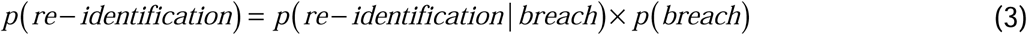

The probability of a breach is computed from published reports on health data breaches and their likelihood that are produced on a regular basis by security companies.

After computing the risk values for the three types of attack, the maximum across them is then taken to reflect the overall risk in the data. If this maximum risk is below the 0.09 threshold, then the dataset is deemed to have an acceptably low risk of re-identification. The same approach is applied to analyse the risk in clinical trial data and in the header information in DICOM files.

#### Strict Average Risk

The risk calculation described above gives us the average risk (averaged across all patients). The strict average conditions this on no records in the dataset being unique in the population. The population is defined as all patients who have participated in clinical trials in the same therapeutic area over the same period and geography. There are a number of estimators that can be used for estimating population uniqueness, with a specific one recommended based on a comparative assessment [16].

#### Application of the Basic Principles

The basic principles have been operationalized for the anonymisation of clinical trial data as a series of default anonymisation practices, which can then be adjusted to account for study-specific data issues. Patient identifiers (typically consisting of a clinical centre number and the patient’s randomization number with which a patient is identified in a clinical trial) is replaced by an anonymised identifier, a new number specifically and uniquely generated for the use of the data in the context of the collaboration. The link file that connects the original patient identifier from the trial with the new anonymised identifier is securely protected and only accessible to a very small independent team who are working exclusively on the anonymisation of the data but who are not otherwise involved in the collaboration or the subsequent research. The link file is used for the sole purpose of assigning the same anonymised patient ID to both the patient’s clinical data and MRI images, so that the corresponding imaging and clinical data remain together after the anonymization is completed, for downstream analyses. By default, event dates in the dataset are offset into relative dates as defined in the PhUSE standard [17]. Also, variables like age are typically generalized to, for example, five year ranges, or modified by adding uniform noise. The SiteID is suppressed so that the geographic location of a site cannot be determined by looking up recruitment information in public registries. Other variables that may contain information that could lead to the re-identification, such as a patient’s medical history, can be generalised or suppressed. The decision as to which variables are transformed takes the intended research purpose into account to preserve the data as much as possible where critical for the research while still bringing the risk of re-identification below the defined threshold. A detailed report is produced documenting the anonymisation methodology, how it was operationalised for each dataset, and a summary of the anonymisation outcomes (e.g., which variables were transformed and how). This detailed report is crucial for the data wrangling and downstream analysis. All data in the final relational database could be linked to the report which described the steps taken.

#### Magnetic Resonance Imaging (MRI) Data

In clinical trials MRI images are commonly obtained at hospitals in Digital Imaging and Communications (DICOM) format and provided to Clinical Research Organisations (CROs) who specialise in imaging analysis. In the MS project the DICOM images were transferred from the CROs to the isolated anonymisation computational environment. Within this environment the image data went into a three-stage process of image conversion, defacing and data curation. Each DICOM file represents one slice of the entire scan; one MRI session will generate multiple scans. The DICOMs were re-assembled into a single 3D volume (as per the research needs), using the DICOM conversion software HeuDiConv [18]. The resulting output of this process is a set of files in a different format (JSON – JavaScript Object Notation, and NIfTI – Neuroimaging Informatics Technology Initiative) that exactly preserve original DICOM data values as well as their associated non-identifying DICOM metadata (i.e. meta-data that could contribute to the identification of patients was stripped out during conversion), but organised in a research ready format, developed and used extensively within the Neuroimaging research community, called Brain Imaging Data Structure (BIDS -https://bids.neuroimaging.io/). In addition to a controlled, standardised file-structure, BIDS provides a file naming convention with the same characteristics, including adding scan-type details for ease of processing. During the initial conversion process, scans that failed to convert, or converted with errors, were put aside and evaluated to see if they could be successfully converted. Overall, we were able to convert scans for over 99% of subjects.

Once the data have been converted to NIfTI and is in BIDS format, they were run through a processing pipeline, simply called ‘defacing’. This pipeline has several steps that aim to achieve two key objectives:

1. Remove identifiable facial features (nose, mouth, front of the eyes, ears)
2. Remove identifiable metadata from the scan’s associated JSON

For privacy/security reasons the identifiable facial features were removed (defaced). The facial identifiable elements were selected according to the anonymisation principles used in the UK BioBank project [19], and removed using defacing software from the FSL software library [20]. To ensure the successful anonymisation, all defacing results are visually checked via multiple 3D surface renderings, confirming the removal of facial features and the retention of brain and meninges. Scans were QC checked and classified as either ‘passed’, or as one of four subclasses of defacing issues. Due to this being a multi-site, longitudinal dataset, MRI scans were of variable quality, and initial defacing failure rates were up to 40% in some studies. Scans that failed QC checks were put through additional rounds of re-defacing and subsequent QC checks, where custom defacing parameters – derived from the type of previous QC classifications and scan modality – were applied to the scans, allowing us to achieve high rates of successful defacing (96%). In total over 230,000 MRIs were defaced and manually checked before entering the research ready dataset. The anonymised data is stripped of all metadata except non-identifiable acquisition parameters. Additional checks were also undertaken to ensure that identifiable details had not been erroneously inserted (during acquisition) into the retained metadata fields. Once all QC checks have been completed, this data is copied via a dedicated and automated mechanism from the anonymisation environment to the analytics environment. Additional safeguards have been implemented to ensure confidentiality and integrity of the data.

### Data Exploration, Quality Control and Integration – Research-Ready Dataset

Non-imaging clinical datasets are anonymised by a third party (Privacy Analytics, Inc) and transferred to the analytics environment at the BDI to begin the data wrangling process. This process has the ultimate goal of providing all data in a relational database, from where streamlined, research ready datasets for the analytics team can be retrieved. A detailed tracking system was developed that is shared across the collaboration to transparently convey the status of each data set within the pipeline. Due to the large number of steps, transformations, and transactions that each dataset goes through, it is essential to track each data point received.

The initial stage in the extract, transform and load (ETL) of data from Novartis to the BDI, was the capture of the clinical trial data as Statistical Analysis Software (SAS) files. For both the MS and IL-17 projects, the BDI team worked closely with the clinical teams at Novartis to ensure full understanding of the data to be downloaded and all related documentation. Each study was downloaded separately in an average of 30 to 50 different tables. These tables contained the primary raw data and study-specific information as described in methods. Each table contained hundreds of thousands of measurements, across hundreds of variables.

Once the data was received, the next critical collaborative step in the process was the data exploration by a dedicated data wrangler to validate that the data received matched the expected data and the protocol documents. This step was extensive and performed in collaboration with the team at Novartis who could review identified queries by exploring the primary data which has not been anonymised. At this stage the tracking of data and related queries between the data generators (Novartis) and the BDI was paramount to ensure all downstream analysis would be reproducible and linkable to a well-defined set of data.

Once the data was agreed to be correct and valid, the data wrangling team developed codebooks (data specifications) which described the structure of the data in a computationally readable format. These codebooks are then utilised by a bespoke software pipeline to homogenise the data into a generic relational database structure. This task had many challenges due to varying trial designs, inconsistencies in data capture between trials, changes in technology throughout history, subjective evaluations, changes in data standards, and anonymisation. Upon successful completion of this part of the pipeline individual data files were imported into the relational database. In conjunction with the import of clinical trial data, the pipeline imports metadata about the relevant additional datasets provided e.g imaging or omics. The key remit for this level of integration is to ensure that relevant data slices can be provided downstream to the analysts and also to ensure the data outputs from the analytics can be integrated back into the overall architecture. This work was deemed to be important, as it was setup in a way that it can be reproduced as new datasets come onboard and was developed in a manner to manage any clinical trial data not just the current data from this project.

Once the data dictionaries were completed across the projects, the data wrangling team began in parallel the overall data quality control process in parallel with the aim of identifying any data quality issues through the data life cycle. The data validation and QC process are innovative as they have been created in a sustainable manner to ensure data is tracked and checked throughout the lifetime of the project and that data provenance is managed at the level of individual data points. The quality of the data from Novartis to the analytical teams was deemed critically important, to ensure data analysts did not have to perform this level of exploratory work and that the results they identified were reproducible. A robust QC pipeline was therefore developed through intensive collaboration that assessed the data at many levels. The quality control pipeline was developed to perform both validation and verification at different stages. For example, source validation ensured the data received matched what was expected and global validation checked the merged data. Validation and verification encompass a large list of checks, from structural checks, assessing levels of missingness, the effects of the anonymisation, and visual checks for potential data anomalies. The anonymisation reports were key to check whether data was missing because it was not captured in the first place, or if it was suppressed due to anonymisation.

The final output of the ETL process was to generate snapshots of data as tracked and versioned data releases for the analysts. A data release is a snapshot of merged datasets available as a relational database or in a data format that can be inputted to analytical tools (e.g. API, table structure etc.). The analytical teams can therefor develop methods which are attributed to the correct version ensuring transparency and reproducibility. As the data analytical methods are being developed, the ETL pipeline is being expanded to ensure that data outputted from new methods can be integrated back into data releases. This is key in a data project.

## Discussion

Novartis and the University of Oxford’s Big Data Institute (BDI) have established a research alliance which has developed an innovative IT platform to manage large volumes of anonymised data. The IT infrastructure that has been developed for this project has enabled the alliance to successfully capture, anonymise, quality control, integrate, and explore data from a large collection of Novartis clinical trials in one research ready environment. This research ready database is now available to a highly multi-disciplinary team of researchers who are analysing and interpreting the data to gain insights about the diseases. The data integrated in this project has not been compiled before, and therefore the technology developed here is allowing data analysts an unprecedented opportunity to develop methods to gain insights across different data modalities (imaging, omics, clinical and biological) and to identify novel patterns with clinical relevance which cannot be detected by humans alone to identify phenotypes, and early predictors of patient disease activity and progression and to improve prognosis for patients. The collaboration currently focuses on Multiple Sclerosis (MS) and other inflammatory diseases in dermatology and rheumatology which are major areas of drug development, but may extend the scope to other disease areas at a later time point.

A milestone achievement of the collaboration is the development of a data anonymisation pipeline for multi-modality data (including clinical and imaging data) which ensures data privacy while preserving the essential clinically relevant pattern in the data. Data anonymization at this scale has the potential to create datasets which are unusable for analysis, so a key step in this project was that specific adjustments were incorporated into the overall anonymisation process that ensured the analytical questions could be addressed. A fundamental component of successful data analysis and the collaborative development of novel machine learning methods on this rich data sets has been the construction of a research informatics framework that can capture the data at regular intervals from Novartis into an IT infrastructure at the Big Data Institute (BDI) where images could be anonymised and integrated with the de-identified clinical data, quality controlled and compiled into a research-ready relational database which would then be available to multi-disciplinary analysts. The collaborative development from a group of software developers, data wranglers, statisticians, clinicians and domain scientists across both organisations has been key. The project has proactively engaged a number of external academic researchers in a number of fields to work with the consortium, get access to the data and contribute to the overall strategic vision. This framework is innovative, as it facilitates collaborative data management and makes a complicated clinical trial data set from a pharmaceutical company available to academic researchers in a secure, granular and robust way. The level of data tracking and data provenance incorporated will ensuring reproducibility and transparency.

## Conclusion

The research alliance has developed an informatics framework to capture multi-dimensional clinical trial data into a pipeline of anonymisation, quality control, data exploration and subsequent integration into a research-ready database. With an emphasis on ensuring data privacy while allowing the development of analytical tools to be conducted, the framework can extend to research on other disease areas, and its principles can be transversally applied into other data settings, especially ones with data privacy concerns.

## Data Availability

Not applicable

## Abbreviations

BDI: Oxford’s Big Data Institute
MS: Multiple sclerosis
PsA: Psoriatic arthritis
RA: Rheumatoid arthritis
PsO: Psoriasis
axSpA: Axial spondyloarthritis
CNS: Central nervous system
RMS: Relapsing multiple sclerosis
CIS: Clinically isolated syndrome
RRMS: Relapsing-remitting multiple sclerosis
SPMS: Secondary progressive multiple sclerosis
PPMS: Primary progressive multiple sclerosis
EDSS: Expanded Disability Status Scale
FLAIR: Fluid-attenuated inversion recovery
MRI: Magnetic Resonance Imaging
DMZ: Demilitarized zone
GDPR: General Data Protection Regulation
EMA: European Medicines Agency
DICOM: Digital Imaging and Communications Format
CROs: Clinical Research Organisations
JSON: JavaScript Object Notation
NIfTI: Neuroimaging Informatics Technology Initiations
BIDS: Brain Imaging Data Structure
ETL: Extract, Transform and Load

## Declarations

### Ethics approval and consent to participate

Not applicable

### Consent for publication

Not applicable

### Availability of data and materials

Not applicable

## Acknowledgements

This collaboration was made possible with the subsequent access to clinical trial data from Novartis. We are grateful for the assistance and facilitation from Prof. Gil McVean for leading the project, Ms. Joanna Stoneham, Mr. Byron Jones and Dr. Anna Zalevski for managing the project, Prof. Lars Fugger, Prof. Mark Jenkinson and Dr. George Nicholson as advisors for data analysis. We are also very grateful to Mr. Henrik Westerberg and Mr. Aaron McCoy for their impactful contribution to key aspects of the MRI anonymisation pipeline; Dr. Bartek Papiez, Dr. Soroosh Afyouni, Dr. Angelos Armen, Dr. Brieuc Lehmann, Mr. Amit Knanna, Mr. Karan Rajesh, Mr. Gordon Graham, Mr. Frank Freischlaeger, Mr. Stephen Robertson and Mr. Costantino Catuogno for data preparation and input on analysis on the MS project; Mr. Shephard Mpofu, Mr. Brian O. Porter, Mr. Hanno Richards, Ms. Luminata Pricop, Ms. Ana de Vera, Ms. Yanli Chang, Ms. Stephanie Danetz, Mr. Michael Beste, Mr. Thibaud Coroller, Mr. Matthias Kormaksson, Ms. Tingting Zhuang, Ms. Xuan Chu, Mr. Albert Widmer, Ms. Ruvie Martin and Mr. Chengeng Tian for data preparation and input on analysis on the IL17 project; Ms. Natasa Hadjistephanou for supporting data wrangling; Mr. Timor Kadir and Mr. Amir Jamaludin for input on spinal MRI and image segmentation; Mr. Robert Esnouf, Mr. Richard Urbanski, Mr. Tim O’Sullivan and Mr. Arnold Ingo for IT infrastructural and security support.

## Authors’ contributions

AMM, DH, SH, KE and TN lead the drafting of the paper, with all other authors helping to contribute to the drafting, reading and final approval of the manuscript. AMM, DD, SG, CHK, DMW, ES and LS contributed to data exploration, data wrangling, quality control, anonymisation of MRI images and development of databases. AH, TH and MCL performed the design, development and maintenance of the IT infrastructure and information security architecture. DH, FD, PA, SA, HG and PK performed data analysis on the MS project. SH, LK, GL and AR performed data analysis on the IL-17 project. KE lead the anonymisation of clinical data. TN, KL, JB and CH oversee and manage both projects within the research collaboration.

